# Deep learning predicts cardiac output from seismocardiographic signals in heart failure

**DOI:** 10.1101/2025.07.11.25331386

**Authors:** Jesse Wang, Seyed M. Nouraie, Neil J. Kelly, Stephen Y. Chan

## Abstract

**Background:** Determination of cardiac output (CO) is essential to the clinical management of cardiovascular compromise. However, the invasiveness, procedural risks, and reliance on specialized infrastructure limit accessibility and scalability of standard-of-care right heart catheterization (RHC). Seismocardiography (SCG), a non-invasive technique which records subtle chest wall vibrations generated by cardiac mechanical activity, may offer a promising alternative for CO determination.

**Objectives:** To develop and evaluate a deep learning model for estimating CO directly from SCG, electrocardiogram (ECG), and body mass index (BMI) in heart failure patients undergoing RHC.

**Methods:** We trained a deep convolutional neural network for CO estimation using an open-access dataset comprising 73 heart failure patients with simultaneous RHC, SCG, and ECG recordings. Model performance was evaluated using a rotating leave-pair-out cross-validation strategy.

**Results:** When estimating CO, the deep learning model achieved a mean bias of -0.35 L/min with limits of agreement (LoA) from -2.21 to 1.51 L/min. When predicting cardiac index in patients with a reference index < 2.2 L/min/m^2^, the model yielded a mean bias of 0.07 L/min/m^2^ with LoA from -0.35 to 0.48 L/min/m^2^.

**Conclusions:** This study demonstrates the feasibility of using deep learning in combination with wearable SCG sensors to non-invasively estimate CO. Model performance was particularly strong in low-output states. These findings highlight the potential of SCG-based monitoring to augment clinical decision-making in settings where invasive measurements are impractical or unavailable. Prospective multicenter validation is needed to confirm generalizability and assess clinical impact.

**Sources of Support:** This work was supported by NIH grants T32 HL129964 (N.J.K.), K08 ES037420 (N.J.K.), R01 HL124021 (S.Y.C.), R01 HL122596 (S.Y.C.); R01 HL151228 (S.Y.C.); the McKamish Family Foundation, the Hemophilia Center of Western Pennsylvania, and the Institute for Transfusion Medicine (N.J.K.), United Therapeutics Jenesis Innovative Research Award (N.J.K.), and the Pulmonary Hypertension Association (N.J.K.).

**Disclosures:** S.Y.C. has served as a consultant for Merck, Janssen, and United Therapeutics; S.Y.C. is a director, officer, and shareholder in Synhale Therapeutics and Amlysion Therapeutics; S.Y.C. and N.J.K. hold research grants from United Therapeutics; S.Y.C. holds research grants from Bayer and the WoodNext Foundation. S.Y.C. has filed patent applications regarding the targeting of metabolism in pulmonary hypertension. Other authors: none.

**Twitter Summary:** We developed a deep learning model to non-invasively estimate cardiac output from wearable seismocardiogram (SCG) signals in patients undergoing right heart catheterization. This is the largest study to date using SCG for noninvasive cardiac output monitoring. #HeartFailure, #WearableTech, #AIInCardiology.

## Introduction

Accurate bedside assessment of cardiac output (CO) is fundamental to the diagnosis and management of cardiovascular compromise and plays a central role in clinical decisions involving inotropic medication, mechanical circulatory support, and treatment escalation^1–3^. Despite the clinical value of CO measurement, its dependence on invasive right heart catheterization (RHC)— which is associated with procedural risks while requiring skilled operators and advanced instrumentation—limits the availability of CO as a clinical tool. In fact, a recent prospective multicenter study reported that cardiogenic shock patients initially evaluated at facilities without invasive monitoring capabilities had higher mortality rates compared to patients who were directly admitted to tertiary care centers^4^. Although echocardiography and magnetic resonance imaging (MRI) can provide certain non-invasive measures of CO, both modalities have limitations: echocardiography relies on geometric assumptions and consistent image quality^5,6^, while MRI may be restricted by high cost and uneven geographic availability^7,8^. These findings underscore the need for accurate, accessible, and non-invasive solutions to expand timely hemodynamic evaluation in settings where RHC is unavailable or contraindicated.

Recent evidence indicates that seismocardiography (SCG) may enable a scalable, non-invasive estimation of key hemodynamic parameters^9–12^. Seismocardiography (SCG) employs multiaxial accelerometers affixed to the chest wall to capture microvibrations of the thoracic surface arising from the mechanical activity of the heart, including myocardial contraction, valvular motion, and blood flow dynamics. Prior studies have suggested the potential of SCG-based models to predict Doppler- and MRI-derived stroke volume (SV) in perioperative and pediatric cohorts^11,12^. However, it is unknown whether SCG can be leveraged for the determination of CO in adults with heart failure.

In this study, we address this gap by evaluating SCG in a publicly available adult heart failure cohort undergoing invasive hemodynamic evaluation^13^. We developed and validated a deep learning model to estimate CO directly using SCG, electrocardiogram (ECG), and body mass index (BMI) as inputs (Central Illustration). All reference CO values were obtained from RHC, the clinical standard of care^3^.

## Methods

### Dataset and code availability

The dataset used in this study is publicly available on the PhysioNet repository at https://physionet.org/content/scg-rhc-wearable-database/1.0.0/^13^. The dataset was collected under a protocol reviewed and approved by the University of California, San Francisco (UCSF) Institutional Review Boards (IRB number: 16-20442) on December 20, 2016. Patients were recruited from the catheterization laboratory at UCSF and all patients provided written consent. The code used for model development and analysis is publicly available at https://github.com/jwang6174/scg-co.

### Study population

The deidentified, open-source dataset released by Chan et al. contained wearable patch signals and RHC measurements for 73 patients referred for hemodynamic evaluation for primary diagnosis of heart failure^11,13^. As nineteen patients had multiple encounters, a total of 83 encounters were included. Each encounter included simultaneously recorded SCG, ECG, and RHC signals. Only baseline hemodynamic parameters before physiologic challenge were analyzed. Patient demographics, clinical characteristics, hemodynamic parameters, and patch signals were collected at the time of RHC. CO was calculated as the average of Fick and thermodilution (TD) when both were available (n = 75 encounters) and was otherwise considered to be the sole Fick (n = 1 encounter) or TD (n = 7 encounters) measurement. Additional details on data preprocessing and signal augmentation are provided in the Supplemental Methods.

### Model design and implementation

Tri-axial SCG signals were used to capture inter-axis mechanical motion and improve robustness to variability in sensor placement. A single-lead ECG, obtained from the wearable patch, was incorporated to provide precise temporal alignment with cardiac events and to enhance model learning of the electromechanical relationship. Prior work has shown improved SV prediction with combined SCG and ECG inputs⁹. BMI was included as an additional input to account for inter-individual differences in signal attenuation related to body habitus^14^. All features were fed into a deep convolutional neural network model. The model was implemented using the PyTorch 2.5.1 programming library^15^. Model training was performed on hardware comprising an Intel Core i7-4790K processor, NVIDIA GeForce GTX 1080 Ti GPU, and 32 GB system memory. Additional details on training and validation strategy are provided in the Supplemental Methods.

### Evaluation metrics and statistical analysis

Root mean squared error (RMSE), relative error, and Pearson correlation coefficient (PCC) were calculated using averaged model predictions for each catheterization encounter. To quantify uncertainty in RMSE, nonparametric bootstrapping was performed by resampling 10,000 bootstrap replicates with replacement from the full set of reference–predicted CO pairs. The 95% confidence interval (CInt) was defined as the 2.5^th^ and 97.5^th^ percentiles of the resulting RMSE distribution. Relative error was expressed as RMSE% and calculated as the RMSE divided by the mean of the reference values then multiplied by 100. For PCC, the 95% CInt was computed via Fisher’s z-transformation, with percentile bounds of the transformed distribution inverse-transformed to the correlation scale. Bland-Altman analysis was conducted to assess agreement between predicted and reference CO values, with mean bias and 95% limits of agreement (LoA). Summary metrics were also calculated across stratified ranges of CO and CI. Statistical analyses were performed with the SciPy 1.14.1 programming library^16^.

## Results

### Demographic and hemodynamic profile of a high-risk heart failure cohort

We first examined patient characteristics to understand how the study population compares with the broader heart failure population. Patient demographic and hemodynamic characteristics were summarized at the encounter level (**Table 1**). The mean age of the cohort was 54.9 ± 13.3 years, and 66.3% of participants were male. The average BMI was 29.2 ± 6.6 kg/m², and the average body surface area (BSA) was 2.0 ± 0.3 m^2^. Among patients in the cohort, 19.3% were NYHA II, 62.7% were NYHA III, 79.5% had heart failure with reduced ejection fraction (HFrEF), and 18.0% had heart failure with preserved ejection fraction (HFpEF). The average heart rate (HR) was 74.3 ± 16.5 bpm. Cardiac rhythms discerned from ECG recordings were 61.8% sinus rhythm, 9.6% atrial fibrillation, 10.8% dual AV paced, and 14.5% V paced.

**Table 1.** Overview of patient characteristics and hemodynamics. Demographic, clinical, and hemodynamic characteristics for all catheterization encounters, validation-test patients, and training-only patients. Values summarized at the encounter level and presented as mean ± SD for normally distributed values and n (%) for counts. AV – atrioventricular; BMI – body mass index; BSA – body surface area; CABG – coronary artery bypass graft; CI – cardiac index; CO – cardiac output; CRT – cardiac resynchronization therapy; EF – ejection fraction; HR – heart rate; ICD – implantable cardioverter defibrillator; mPAP – mean pulmonary artery pressure; NYHA – New York Heart Association; OHTx – orthotic heart transplant; PAC – premature atrial contraction; PAWP – pulmonary artery wedge pressure; PCI – percutaneous coronary intervention; PVC – premature ventricular contraction; RAP – right atrial pressure; SV – stroke volume.

|  | **All**<br>**(n = 83)** | **Validation-Test**<br>**Set (n = 64)** | **Training-Only Set**<br>**(n = 19)** |
| --- | --- | --- | --- |
| **Age (years)** | 54.9 ± 13.3 | 55.9 ± 13.8 | 51.6 ± 10.9 |
| **Sex** |  |  |  |
| **Male** | 55 (66.3%) | 45 (70.3%) | 10 (52.6%) |
| **Female** | 28 (33.7%) | 19 (29.7%) | 9 (47.4%) |
| **BMI (kg/m<sup>2</sup>)** | 29.2 ± 6.6 | 29.9 ± 7.0 | 26.9 ± 4.2 |
| **BSA (m<sup>2</sup>)** | 2.0 ± 0.3 | 2.1 ± 0.3 | 1.9 ± 0.2 |
| **NYHA** |  |  |  |
| **I** | 6 (7.2%) | 6 (9.4%) | 0 (0%) |
| **II** | 16 (19.3%) | 14 (21.9%) | 2 (10.5%) |
| **III** | 52 (62.7%) | 40 (62.5%) | 12 (63.2%) |
| **IV** | 6 (7.2%) | 4 (6.2%) | 2 (10.5%) |
| **Not Reported** | 3 (3.6%) | 0 (0%) | 3 (15.8%) |
| **Outpatient** |  |  |  |
| **Yes** | 67 (80.7%) | 52 (81.2%) | 15 (78.9%) |
| **No** | 16 (19.3%) | 12 (18.8%) | 4 (21.1%) |
| **EF** |  |  |  |
| **Preserved** | 15 (18.0%) | 11 (17.2%) | 4 (21.1%) |
| **Reduced** | 66 (79.5%) | 51 (79.7%) | 15 (78.9%) |
| **Not Reported** | 2 (2.4%) | 2 (3.1%) | 0 (0%) |
| **Other History** |  |  |  |
| **CABG** | 10 (12.0%) | 10 (15.6%) | 0 (0%) |
| **CRT-D** | 24 (28.9%) | 20 (31.2%) | 4 (21.1%) |
| **ICD** | 25 (30.1%) | 19 (29.7%) | 6 (31.6%) |
| **OHTx** | 6 (7.2%) | 2 (3.1%) | 4 (21.1%) |
| **PCI** | 9 (10.8%) | 7 (10.9%) | 2 (10.5%) |
| **VAD** | 2 (2.4%) | 0 (0%) | 2 (10.5%) |
| **RAP (mmHg)** | 8.2 ± 5.3 | 8.7 ± 5.4 | 6.3 ± 4.8 |
| **mPAP (mmHg)** | 28.6 ± 10.6 | 30.1 ± 10.9 | 23.3 ± 7.5 |
| **PAWP (mmHg)** | 17.1 ± 7.7 | 17.7 ± 8.0 | 14.7 ± 6.1 |
| **HR (bpm)** | 74.3 ± 16.5 | 74.4 ± 15.8 | 74.1 ± 19.2 |
| **Rhythm** |  |  |  |
| **Sinus Rhythm** | 51 (61.4%) | 39 (60.9%) | 12 (63.2%) |
| **Atrial Fibrillation/Flutter** | 8 (9.6%) | 6 (9.4%) | 2 (10.5%) |
| **Paced Rhythm** | 24 (28.9%) | 19 (29.7%) | 5 (26.3%) |
| **A-Paced** | 1 (1.2%) | 1 (1.6%) | 0 (0%) |
| **AV-Paced** | 9 (10.8%) | 7 (10.9%) | 2 (10.5%) |
| **V-Paced** | 14 (16.9%) | 11 (17.2%) | 3 (5.3%) |
| **SV (mL)** | 64.8 ± 20.7 | 64.6 ± 21.1 | 65.6 ± 19.7 |
| **CO (L/min)** | 4.7 ± 1.7 | 4.6 ± 1.4 | 5.0 ± 2.4 |
| **CI (L/min/m<sup>2</sup>)** | 2.3 ± 0.9 | 2.3 ± 0.6 | 2.6 ± 1.5 |

**Table 2.** Summary of model performance metrics by encounter subgroup. Units were omitted for visual clarity. CI was represented in L/min/m^2^, and CO was represented in L/min. All PCC values demonstrated statistical significance of P < 0.005. CI – cardiac index; CO – cardiac output; LoA – limits of agreement; PCC – Pearson correlation coefficient; RMSE – root mean squared error.

|  | Subgroup (n) | RMSE<br>(95% CInt) | RMSE% | PCC<br>(95% CInt) | Mean Bias<br>(95% LoA) |
| --- | --- | --- | --- | --- | --- |
| **CO** | All (64) | 1.00<br>(0.69-1.30) | 22% | 0.75<br>(0.61-0.84) | -0.35<br>(-2.21 to 1.51) |
|  | CO < 6 (53) | 0.44<br>(0.34-0.54) | 12% | 0.80<br>(0.68-0.88) | -0.01<br>(-0.88 to 0.87) |
|  | CI < 2.2 (36) | 0.42<br>(0.30-0.54) | 11% | 0.76<br>(0.58-0.87) | 0.12<br>(-0.68 to 0.92) |
|  | CI < 1.8 (15) | 0.41<br>(0.25-0.59) | 12% | 0.77<br>(0.42-0.92) | 0.19<br>(-0.55 to 0.93) |
| **CI** | All (64) | 0.46<br>(0.32-0.58) | 20% | 0.71<br>(0.56-0.81) | -0.15<br>(-1.01 to 0.70) |
| | CO < 6 (53) | 0.23<br>(0.17-0.29) | 11% | 0.81<br>(0.70-0.88) | $1.0 \times 10^{-3}$<br>(-0.45 to 0.45) |
|  | CI < 2.2 (36) | 0.22<br>(0.14-0.29) | 12% | 0.73<br>(0.53-0.85) | 0.07<br>(-0.35 to 0.48) |
|  | CI < 1.8 (15) | 0.20<br>(0.12-0.28) | 12% | 0.73<br>(0.34-0.90) | 0.09<br>(-0.26 to 0.44) |

Participants had an average CO of 4.7 ± 1.7 L/min and an average CI of 2.3 ± 0.9 L/min/m^2^. The distributions of CO and CI values were approximately normal (**Figure 1A** and **Figure 1B**). An outlier, defined as greater than three standard deviations from the mean, was observed with Fick CO of 9.4 L/min and TD CO of 17.8 L/min, resulting in an average CO of 13.6 L/min. For encounters where both measurements were performed, Fick and TD were strongly correlated with a PCC of 0.84 (95% CInt: 0.76–0.90, P < 0.001, **Figure 1C**). Two RHC encounters were performed while the patient was supported by a ventricular assistive device (VAD) and were both included in the training-only set. Overall, the cohort reflected a high-risk heart failure population characterized by advanced functional limitation, a substantial burden of pacing dependence, and reduced CI. *Deep learning model architecture for predicting CO from SCG signals*. We then developed a deep learning model comprised of parallel convolutional subnetworks for SCG and ECG signals, followed by a fully connected fusion module that integrated these features with BMI (**Figure 2**). Each signal branch applied a sequence of four 1D convolutional layers with a fixed kernel size and increasing dilation factors, allowing the network to capture both local and long-range temporal patterns. The resulting feature maps were collapsed into a single time-step via an adaptive average-pooling layer, then passed through a lightweight residual projection mechanism to provide the model with flexible feature scaling and normalization.

**Figure 1.**
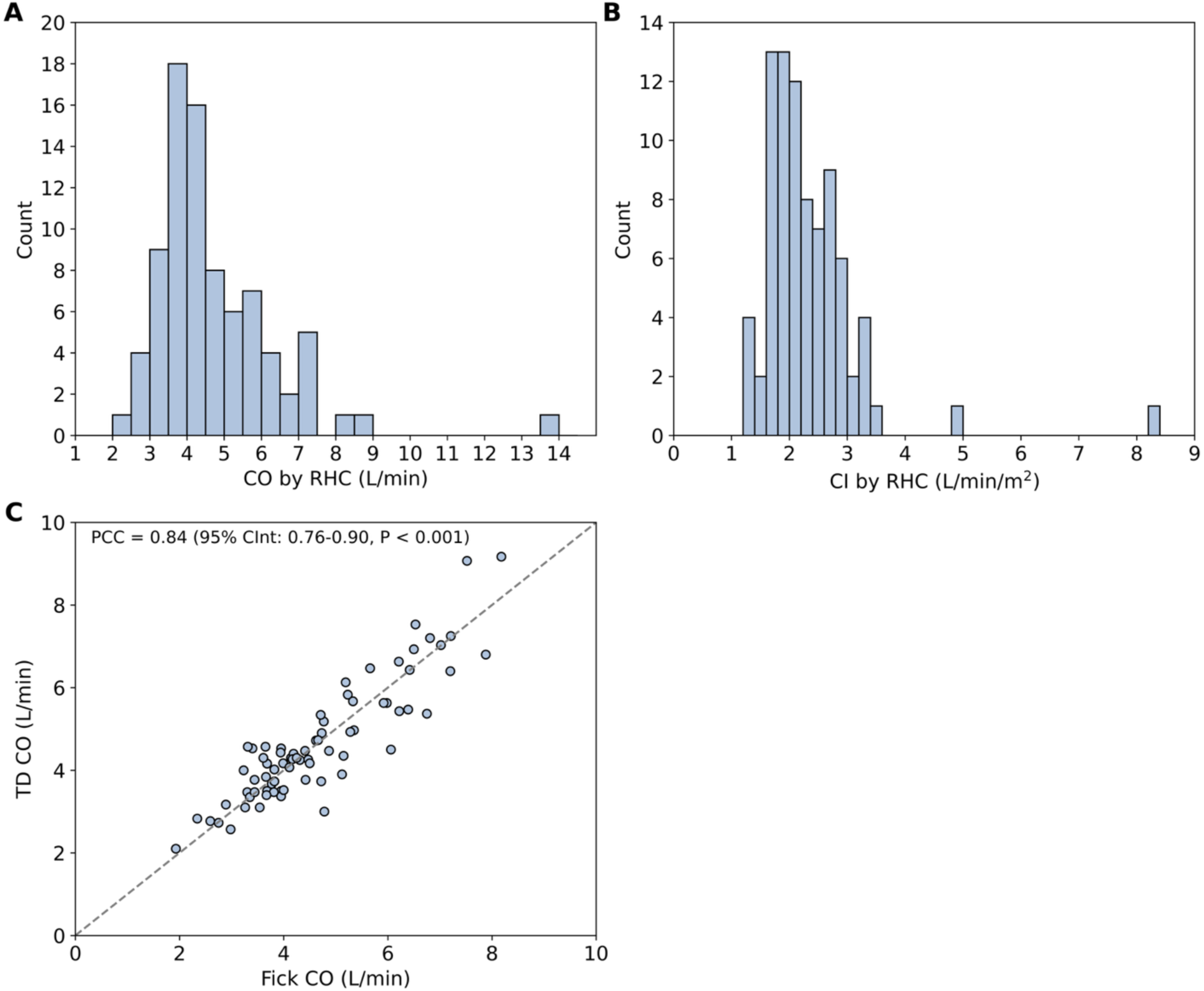
Distribution of CO and CI for all baseline RHC encounters. **(A)** Histogram of CO values as average of Fick and TD obtained during RHC. **(B)** Histogram of CI values calculated as average CO by Fick and TD normalized to BSA. **(C)** Correlation between Fick CO and TD CO. An outlier with Fick CO of 9.4 L/min and TD CO of 17.8 L/min was omitted for visual clarity. BSA – body surface area; CO – cardiac output; CI – cardiac index; CInt – confidence interval; PCC – Pearson correlation coefficient; RHC – right heart catheterization; TD – thermodilution.

**Figure 2.**
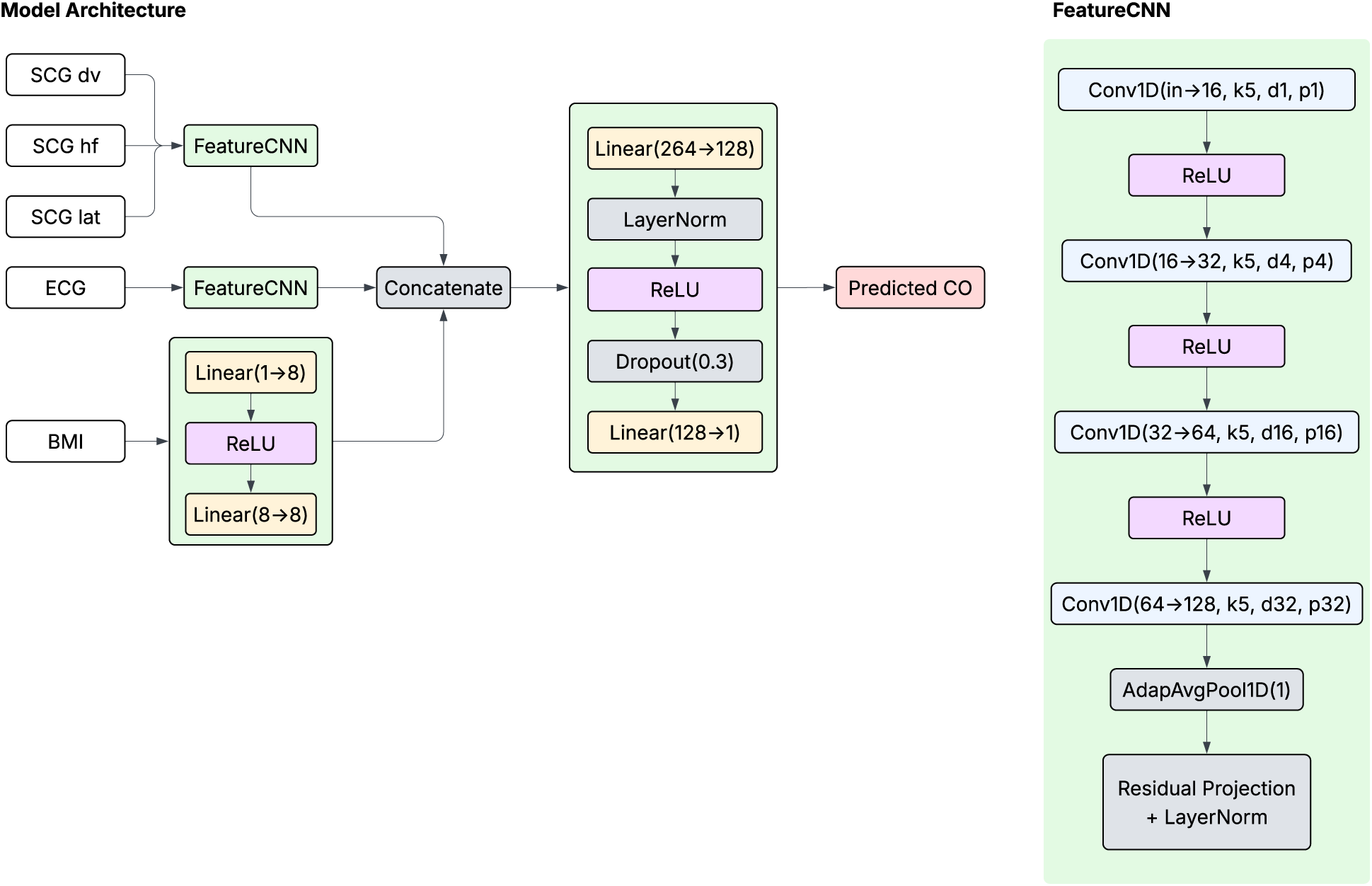
Schematic overview of the deep learning model for CO prediction. Tri-axial SCG and single-lead ECG serve as inputs to parallel convolutional subnetworks (FeatureCNN). A linear layer refers to a fully connected layer that maps a specified number of inputs to a specified number of output units. Dropout, with a given probability *p*, randomly sets a proportion of activations to zero during training to limit overfitting. A 1D convolutional layer (Conv1D) applies a set number of filters of specified kernel size along the temporal axis, with configurable dilation and padding settings to control the receptive field and output size. Adaptive average pooling (AdapAvgPool1D) performs averaging over the temporal axis to produce a fixed-length output, regardless of the input size. BMI – body mass index; CO – cardiac output; ECG – electrocardiogram; SCG – seismocardiogram.

The refined 128D feature representations from the SCG and ECG branches were concatenated with an 8D embedding of BMI, derived from a 2-layer fully connected network. These features were passed through a final multilayer perceptron, consisting of layer normalization, dropout, and a linear output, to predict CO as a continuous variable. All hidden layers used ReLU activation. The network was trained end-to-end using the Adam optimizer with an initial learning rate of 1 × 10^−3^, using a cosine annealing learning rate schedule with warm restarts and a batch size of 128. Mean squared error was used as the loss function.

### Performance of the deep learning model for CO prediction

After model development, we assessed the performance of the model for all patients in the validation-test set. RMSE for the CO predicted by SCG with deep learning compared to the reference CO measured by RHC was 1.00 L/min (95% CInt: 0.69–1.30), RMSE% was 22%, and PCC was 0.75 (95% CInt: 0.61–0.84, P < 0.001, **Figure 3A,B**). The maximum width of the 95% confidence intervals for encounter-level CO values, which were averaged from overlapping 30-second segment predictions, was 0.16 L/min. Bland-Altman analysis demonstrated a mean bias of -0.35 L/min between predicted and reference CO values, with 95% LoA from -2.21 to 1.51 L/min (**Figure 3C,D**). The model demonstrated consistent predictive performance across a range of cardiac rhythms (**Figure 3B,D**). In the validation-test subset of 53 patients with CO < 6 L/min (82.8% of the cohort), RMSE decreased to 0.44 L/min (95% CInt: 0.34–0.54), PCC% decreased to 12%, and PCC increased to 0.80 (95% CInt: 0.68–0.88, P < 0.001, **Table 3**). Mean bias narrowed to -0.01 L/min, with 95% LoA from -0.88 to 0.87 L/min.

**Figure 3.**
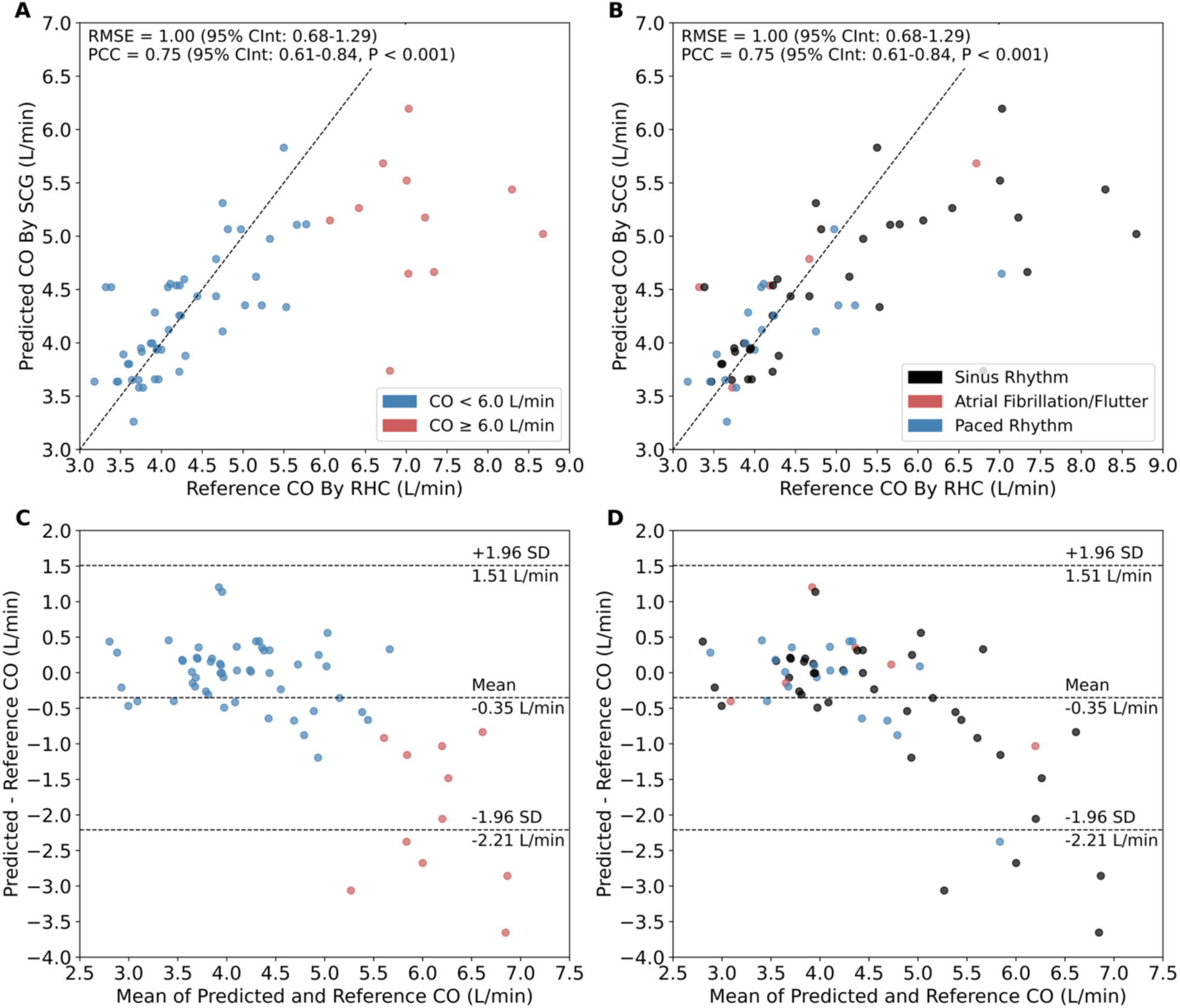
Performance of SCG-based deep learning model for CO prediction. Correlation of predicted CO by SCG versus reference CO by RHC for the validation-test set of 64 patients, color-coded by (**A**) CO and (**B**) cardiac rhythm. Each point represents the averaged predicted CO for all 30-second segments in a catheterization encounter. Error bars were omitted for visual clarity. Maximum width of 95% confidence intervals for all points was 0.156 L/min. Bland-Altman plot between predicted and reference CO values color-coded by (**C**) cardiac output and (**D**) cardiac rhythm. Middle dashed line denotes the mean bias. Upper and lower dashed lines represent the 95% LoA. CInt – confidence interval; CO – cardiac output; LoA – limits of agreement; PCC – Pearson correlation coefficient; RHC – right heart catheterization; RMSE – root mean squared error; SCG – seismocardiogram.

Given the decreased model performance when including 11 patients with CO ≥ 6 L/min (17.2% of the validation-test subset), we performed a targeted analysis of contributing factors. Patients in this CO range exhibited modestly greater BSA compared to patients with CO < 6 L/min (2.3 ± 0.3 vs 2.0 ± 0.3 m^2^; P = 0.021; **Table S1**). However, BMI was not significantly different between the two groups (32.8 ± 6.0 vs 29.4 ± 7.1 kg/m^2^ for CO ≥ 6 and CO < 6 L/min, respectively; P = 0.114). Patients with CO ≥ 6 L/min and those with CO < 6 L/min were of approximately similar age (51.3 ± 15.5 vs 56.8 ± 13.4 years; P = 0.288) and had comparable male predominance (63.6% vs 71.7%; P = 0.719). NYHA class III symptom burden was comparable between groups (54.5% vs 64.2% for CO ≥ 6 L/min and CO < 6 L/min, respectively; P = 0.734). Intracardiac filling pressures were expectedly lower in patients with CO ≥ 6 L/min; pulmonary arterial wedge pressure (PAWP) averaged 13.5 ± 6.7 mmHg in the CO ≥ 6 L/min group compared to 18.6 ± 8.0 mmHg in the CO < 6 L/min group (P = 0.041).

### Performance of the deep learning model for CI prediction

The BSA-normalized CO, or CI, may offer more meaningful information in clinical settings^17^. Therefore, to understand model behavior in terms of CI, we normalized model-predicted CO to BSA and compared it to the reference CI, which was derived by normalizing the average of Fick and TD output measurements to BSA. Between predicted and reference CI values, RMSE was 0.46 L/min/m^2^ (95% CInt: 0.32–0.58), RMSE% was 20%, and PCC was 0.71 (95% CInt: 0.56–0.81, P < 0.001, **Figure 4A,B**). Bland-Altman analysis demonstrated a mean bias of -0.15 L/min/m^2^, with 95% LoA from -1.01 to 0.70 L/min/m^2^ (**Figure 4C,D**). After excluding patients with reference CO ≥ 6 L/min—whose signal characteristics may not have been adequately represented for the model to learn—RMSE decreased to 0.23 L/min/m^2^ (95% CInt: 0.17–0.29), RMSE% decreased to 11%, and PCC increased to 0.81 (95% CInt: 0.70–0.88, P < 0.001, **Table 3**). Mean bias narrowed to 1.0 × 10^−3^ L/min/m^2^, with 95% LoA from -0.45 to 0.45 L/min/m^2^.

**Figure 4.**
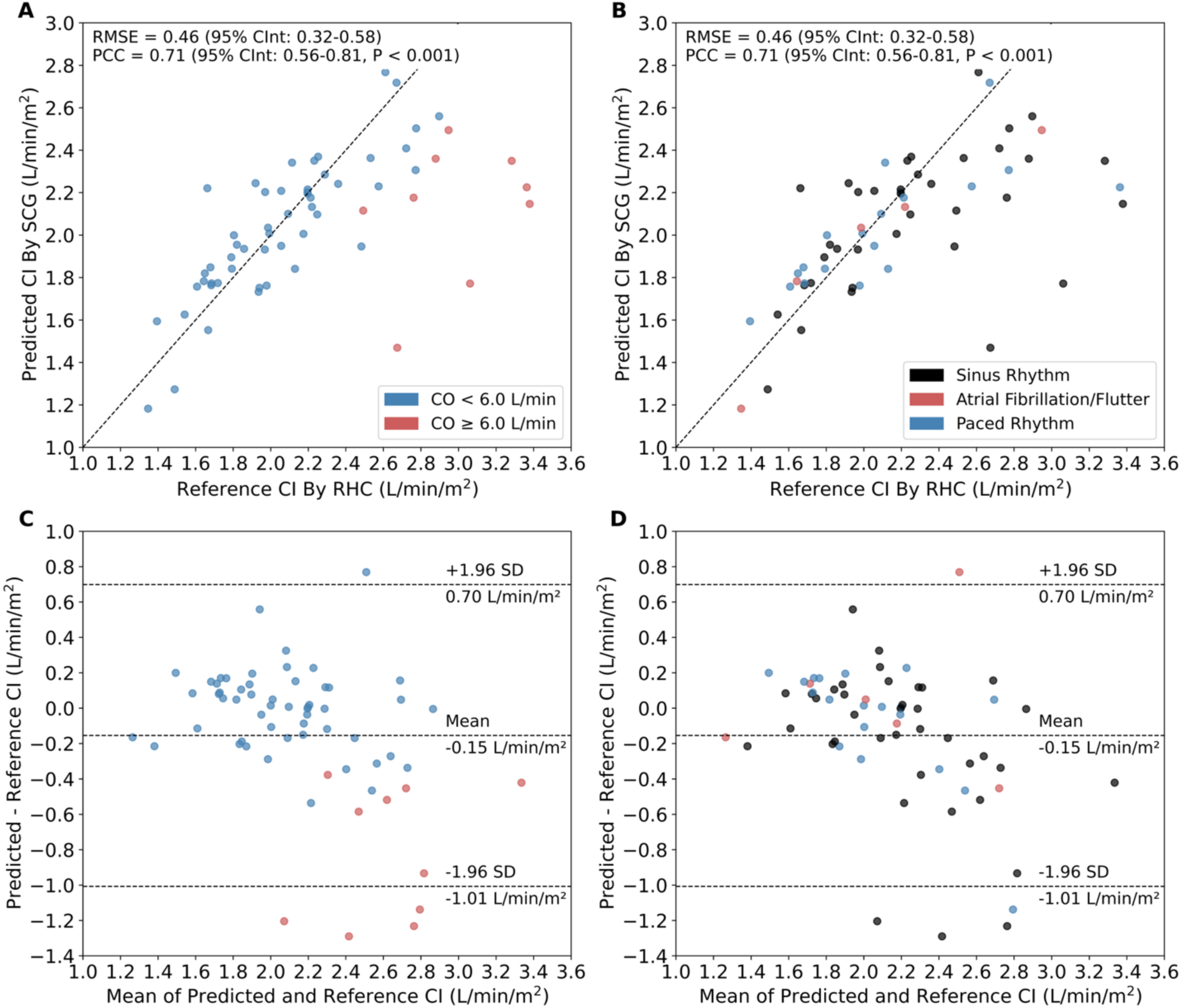
Performance of SCG-based deep learning model for CI prediction. Correlation of predicted CI by SCG versus reference CI by RHC for validation-test set of 64 patients color-coded by (**A**) CO and (**B**) cardiac rhythm. Each point represents the averaged predicted CI for all 30-second segments in a catheterization recording. Error bars were omitted for visual clarity. Maximum width of 95% confidence intervals for all points was 0.062 L/min/m^2^. Bland-Altman plot between predicted and reference CI values color-coded by (**C**) CO and (**D**) cardiac rhythm. Middle dashed line denotes the mean bias. Upper and lower dashed lines represent the 95% LoA. CI – cardiac index; LoA – limits of agreement; PCC – Pearson correlation coefficient; RHC – right heart catheterization; RMSE – root mean squared error; SCG – seismocardiogram.

### Model performance in low-flow hemodynamic conditions

We were next interested in evaluating model performance for CI estimation in low-flow states, which may be of particular clinical relevance^17^. In a subset of 15 patients with CI < 1.8 L/min/m^2^, a threshold suggestive of cardiogenic shock in patients not receiving pharmacologic or mechanical support^17^, RMSE was 0.20 L/min/m^2^ (95% CInt: 0.12–0.28), RMSE% was 12%, PCC was 0.73 (95% CInt: 0.34–0.90, P = 0.002), and mean bias was 0.09 L/min/m^2^, with 95% LoA from -0.26 to 0.44 L/min/m^2^. In a subset of 36 patients with CI < 2.2 L/min/m^2^, a threshold compatible with cardiogenic shock with support^17^, RMSE was 0.22 L/min/m^2^ (95% CInt: 0.14–0.29), RMSE% was 12%, PCC was 0.73 (95% CInt: 0.53–0.85, P < 0.001), and mean bias was 0.07 L/min/m^2^ with 95% LoA from -0.35 to 0.48 L/min/m^2^.

## Discussion

In this study, we developed and validated a deep learning model to noninvasively estimate CO using SCG and ECG signals in patients undergoing RHC. The model demonstrated strong overall performance, with particularly improved accuracy in patients with low CO or CI—ranges of greatest clinical concern in heart failure management. These findings suggest that SCG-based approaches may offer a viable, noninvasive method for estimating hemodynamics in settings where invasive monitoring is impractical or unavailable.

Prediction accuracy was greatest among patients with reduced CI, including those meeting common thresholds associated with cardiogenic shock^17^. The model demonstrated low RMSE and strong correlation to reference CI values. These findings suggest that SCG-derived estimates may be sensitive enough to detect hemodynamic compromise and could potentially support earlier identification and management of patients with impaired perfusion. Together, these results underscore the potential clinical utility of SCG-based deep learning models for noninvasive cardiovascular monitoring.

Model performance declined in patients with reference CO ≥ 6 L/min, a minority subset of the study population. These higher-output encounters were associated with increased residual error and proportional bias on Bland-Altman analysis. The reduced accuracy may reflect limited waveform representation at the higher end of the hemodynamic spectrum, constraining the model’s ability to learn distinguishing features in this range. Notably, patients with elevated CO exhibited slightly greater BSA, which may introduce mechanical filtering or signal scaling effects that confound SCG-based interpretations. Despite this, the majority of patients with CO ≥ 6 L/min were in normal sinus rhythm, suggesting that rhythm abnormalities were unlikely to account for the observed decline in performance.

This limitation in performance at higher cardiac outputs is consistent with prior findings. For example, Semiz et al. reported decreased accuracy when estimating SV > 100 mL, particularly above 150 mL, in a cohort of 12 perioperative patients^10^. They attributed this limitation to the fact that higher SV values came from a single subject, suggesting that limited exposure to higher-output examples during training constrained the model’s generalizability. After excluding patients with CO ≥ 6 L/min in the present study, the model’s mean bias approached zero, and its LoA narrowed considerably, supporting its robustness within the lower-output range most relevant to heart failure and cardiogenic shock populations. Addressing this limitation may involve targeted oversampling, dedicated model pathways for supraphysiologic outputs, or the inclusion of additional training data spanning a broader hemodynamic spectrum.

Compared to earlier efforts such as that by Ganti et al., which employed ridge regression to estimate SV from a single-channel SCG and ECG signals in a cohort of 45 congenital heart disease patients^12^, the present study leverages a deep learning architecture that integrates tri-axial SCG, ECG, and BMI to directly estimate CO. This approach offers several advantages. First, the use of tri-axial SCG enables the model to capture motion across multiple dimensions, enhancing robustness to sensor placement variability and improving sensitivity to complex mechanical patterns such as ventricular heave^18^. Second, by predicting CO directly rather than estimating SV and multiplying by HR, the model may learn an integrated representation of flow dynamics, potentially improving robustness and generalizability across diverse patient states and cardiac rhythms. This is particularly relevant in heart failure, where abnormal rhythms such as atrial fibrillation are common and SV can vary substantially from beat to beat, potentially introducing instability in SV-based approaches^19,20^. Notably, the model demonstrated consistent performance across a range of cardiac rhythms. Finally, deep learning enables the extraction of non-linear, high-dimensional representations that ridge regression cannot capture, particularly in the presence of heterogeneous signal quality and physiological variability^21^. These advantages collectively represent a more comprehensive and scalable framework for non-invasive hemodynamic assessment with SCG.

Several other recent studies have explored the feasibility of using SCG-based models to estimate hemodynamic parameters. For example, Shandhi et al. analyzed SCG signals in 20 heart failure patients and developed a population regression model capable of estimating changes in mPAP and PAWP, with RMSEs of 2.5 mmHg and 1.9 mmHg, and R² values of 0.83 and 0.93, respectively^11^. Additionally, a recent multicenter prospective trial by Klein et al. using a SCG-based deep learning model to estimate PAWP in 310 patients with HFrEF showed an error of 1.04 ± 5.57 mmHg with LoA of -9.9 to 11.9 mmHg^22^. The present study represents, to the best of our knowledge, the first application of SCG-based deep learning for direct CO estimation in adult heart failure patients.

Other modalities, including pulsed-wave Doppler echocardiography using left ventricular outflow tract (LVOT) velocity-time integral and HR, offer alternatives for non-invasively estimating CO^5,6^. This approach, however, relies on accurate alignment of the ultrasound beam with flow and makes assumptions of LVOT geometry. The approach is also operator-dependent and may be limited by suboptimal acoustic windows. Cardiac MRI offers high-fidelity CO estimation but is costly, time-intensive, and restricted to specialized centers^7,8^. Both echocardiography and MRI are limited to discrete snapshots of CO, reducing their utility for continuous or frequent assessment. In contrast, SCG can be acquired continuously via low-profile wearable sensors without the need for imaging expertise, offering the potential for real-time, automated hemodynamic monitoring outside of specialized care settings. These advantages suggest that SCG may be well suited for deployment in resource-limited environments and as a complementary tool for serial monitoring in both inpatient and outpatient settings.

We acknowledge limitations to this study. Study data was derived from a small, single-center, open-access repository, and although it included diverse patient characteristics, it may not fully reflect the heterogeneity of patients across different clinical settings. Due to this constraint, a fixed hold-out set was not employed; instead, model performance was evaluated using LPOCV. Moreover, the study cohort differed from the general adult heart failure population in several aspects. First, the cohort was predominantly male, whereas heart failure affects men and women at similar rates in the broader population^23^. Second, while the general male heart failure population shows a roughly equal distribution of HFpEF and HFrEF, most cases in the study cohort were HFrEF^23^. Third, the study cohort exhibited more advanced disease severity, with a greater proportion of patients classified as NYHA III, compared to the predominance of class II symptoms in the general heart failure population^24^. Although these differences may limit generalizability, the inclusion of higher-risk patients may enhance the clinical relevance of the findings for populations most vulnerable to hemodynamic instability, where accurate noninvasive monitoring is particularly valuable.

## Conclusion

This study demonstrates the feasibility of using SCG and ECG signals to non-invasively estimate CO through a deep learning model. The model achieved strong agreement with catheter-derived values, particularly within the lower-output range relevant to patients with heart failure and cardiogenic shock. While prospective external validation is required, especially in diverse clinical settings and among patients with high-output physiology, these findings highlight the potential of SCG-based monitoring to support scalable, continuous hemodynamic assessment. Unlike MRI or echocardiography—which require specialized equipment, technical expertise, and are limited to intermittent measurements—SCG can be implemented using compact, wearable sensors that enable automated, repeatable, and continuous monitoring. With additional development, such tools may offer a practical, low-risk alternative for bedside evaluation on general medical floors, in outpatient settings, and in resource-limited environments where invasive monitoring is not readily available.

## Clinical Perspectives

### Competency in Medical Knowledge

Accurate, real-time cardiac output assessment plays a vital role in the diagnosis and management of cardiogenic shock and heart failure. This study demonstrates that deep learning applied to seismocardiographic and electrocardiographic signals can provide noninvasive estimates of cardiac output that correlate well with right heart catheterization measurements. Such models may eventually enable bedside hemodynamic monitoring on inpatient wards and in resource-limited settings, offering a scalable and lower-risk alternative to invasive catheterization in select patients.

### Translational Outlook

Further validation in external, multicenter cohorts is needed to confirm generalizability across diverse patient populations and clinical environments. Prospective studies assessing model performance under real-world conditions—including variable sensor placement, ambulatory movement, and pharmacologic interventions—will be essential. Integration into clinical workflows and regulatory considerations must also be addressed prior to clinical adoption.

## Supporting information

Central Illustration

## Data Availability

The code used for model development and analysis is publicly available at https://github.com/jwang6174/scg-co.

## Abbreviations

CI: Cardiac index
CInt: Confidence interval
CO: Cardiac output
LoA: Limits of agreement
LPOCV: Leave-pair-out cross-validation
PCC: Pearson correlation coefficient
RHC: Right heart catheterization
RMSE: Root mean squared error
SCG: Seismocardiogram
TD: Thermodilution

## Acknowledgements

Created in BioRender. Kelly, N. (2025) https://BioRender.com/ylcuuh2.

