## Supplemental Material for "Deep learning predicts cardiac output from seismocardiographic signals in heart failure"

**Supplemental Methods**

*ECG review.* Cardiac rhythms were reviewed by two independent physicians including one board-certified in cardiovascular disease by the American Board of Internal Medicine. Rhythm determination was made based upon the first 10 seconds of the ECG recording; additional frames were reviewed when the rhythm was unclear (e.g. artifactual noise in the initial 10 seconds). Both physicians had access to the publicly available clinical histories during ECG review.

*Data preprocessing and signal augmentation.* Recordings were segmented into 30-second windows with a 3-second overlap. Each segment included tri-axial SCG signals, a single-lead ECG signal, BMI, and a reference CO value measured by RHC. Segments were grouped into CO bins at intervals of 500 mL/min. Random oversampling was applied such that each bin contained approximately equal number of segments as the most populated bin. This strategy was implemented to mitigate class imbalance and promote robust feature learning across the full range of CO values.

To enhance model generalizability and reflect real-world signal variability, several data augmentations were applied during training. These included random temporal shifting of both SCG and ECG signals, addition of Gaussian noise to simulate sensor-related artifacts, and stochastic dropout of SCG channels to mimic partial sensor failure. Physiological parameters were also perturbed: BMI values were randomly varied by 1–4%, and CO labels by 1–2%. These augmentations were applied exclusively during training. No transformations were used during validation or testing.

*Training and validation strategy.* To balance methodological rigor with practical constraints related to computational time and limited sample size, model performance was assessed using an encounter-level leave-pair-out cross-validation (LPOCV) scheme. In each fold, two encounters were withheld: one assigned to validation to guide epoch selection, and the other reserved for final testing. The model was trained on the remaining encounters, with no overlap in data between training, validation, and test sets. After initial evaluation, the roles of the validation and test encounters were reversed within the same training run, and model performance was reassessed without retraining or hyperparameter adjustment. This rotation allowed for two independent test evaluations per fold while preserving a strict patient-wise separation. Ten patients were used for validation in multiple LPOCV folds. To avoid duplication in aggregate performance metrics, each encounter's first appearance as a test case was used. This strategy ensured robust evaluation of model generalizability to unseen encounters, minimizing the risk of data leakage and overfitting.

To maximize available data for training and avoid data leakage, only patients who underwent a single RHC were considered for validation and testing, which included 65 patients. Among these 65 patients, 64 were selected into a validation-test set and were paired to another patient with a similar reference CO, using the following stratification scheme: CI $< 2.2$ L/min/m$^2$, CI $\geq 2.2$ L/min/m$^2$ with CO $< 6$ L/min, and CO $\geq 6$ L/min. One outlier, defined as more than three standard deviations from the mean with a CO of 13.6 L/min was excluded from the validation-test set due to the lack of a comparable CO, although it was included for model training. Stratified pairing ensured that validation and test cases within each fold had similar reference CO values. This approach reduced variability between held-out samples and supported consistent model selection based on validation loss trajectories. Patients in the validation-test set, when not used for validation or testing in an LPOCV fold, were used for model training. Nineteen patients underwent multiple RHCs and were only used for model training, in combination with 62 out of 64 patients from the validation-test set. The other two patients in the validation-test set were used for validation and testing. In summary, in each LPOCV fold, 81 RHC encounters were used for training, one encounter was used as validation to select the optimal number of epochs, and one encounter was used for final model evaluation for that LPOCV fold. Thirty-eight models were trained and their performance on 64 patients were aggregated for analysis.

Model checkpoint selection was based on the epoch corresponding to the lowest validation loss. In instances where the validation loss trajectory demonstrated instability or noise, selection incorporated qualitative review of the loss curve to identify a checkpoint preceding sustained increased in loss. A formal early stopping criteria was not applied; training was discontinued when further epochs failed to yield consistent validation improvement. Checkpoint selection was determined exclusively using the validation patient in each fold. For each catheterization encounter, the final predicted CO was calculated as the mean of the model's predictions for all 30-second segments. Model performance was evaluated by aggregating results across all folds of the LPOCV framework.

**Table S1.** *Overview of patient characteristics and hemodynamics in the validation-test set stratified by CO threshold of 6 L/min.* Values summarized at the encounter level and presented as mean ± SD for normally distributed values and n (%) for counts. Statistical significance between groups was computed using Welch's t-test for continuous variables and Fisher's exact test for categorical variables. AV – atrioventricular; BMI – body mass index; BSA – body surface area; CABG – coronary artery bypass graft; CI – cardiac index; CO – cardiac output; CRT – cardiac resynchronization therapy; EF – ejection fraction; HR – heart rate; ICD – implantable cardioverter defibrillator; mPAP – mean pulmonary artery pressure; NYHA – New York Heart Association; OHTx – orthotic heart transplant; PAC – premature atrial contraction; PAWP – pulmonary artery wedge pressure; PCI – percutaneous coronary intervention; PVC – premature ventricular contraction; RAP – right atrial pressure; SV – stroke volume.

| | **CO < 6 L/min (n = 53)** | **CO ≥ 6 L/min (n = 11)** | **P-value** |
|---|---|---|---|
| **Age (years)** | 56.8 ± 13.4 | 51.3 ± 15.5 | 0.288 |
| **Sex** | | | |
| **Male** | 38 (71.7%) | 7 (63.6%) | 0.719 |
| **Female** | 15 (28.3%) | 4 (36.4%) | – |
| **BMI (kg/m$^2$)** | 29.4 ± 7.1 | 32.8 ± 6.0 | 0.114 |
| **BSA (m$^2$)** | 2.0 ± 0.3 | 2.3 ± 0.3 | 0.021 |
| **NYHA** | | | |
| **I** | 3 (5.7%) | 3 (27.3%) | 0.058 |
| **II** | 13 (24.5%) | 1 (9.1%) | 0.431 |
| **III** | 34 (64.2%) | 6 (54.5%) | 0.734 |
| **IV** | 3 (5.7%) | 1 (9.1%) | 0.539 |
| **Outpatient** | | | |
| **Yes** | 43 (81.1%) | 9 (81.8%) | 1.000 |
| **No** | 10 (18.9%) | 2 (18.2%) | – |
| **EF** | | | |
| **Preserved** | 8 (15.1%) | 3 (27.3%) | 0.384 |
| **Reduced** | 44 (83.0%) | 7 (63.6%) | 0.213 |
| **Not Reported** | 1 (1.9%) | 1 (9.1%) | 0.316 |
| **Other History** | | | |
| **CABG** | 9 (17.0%) | 1 (9.1%) | 1.000 |
| **CRT-D** | 17 (32.1%) | 3 (27.3%) | 1.000 |

| | | | |
|---|---|---|---|
| **ICD**<br>**OHTx**<br>**PCI** | 17 (32.1%)<br>1 (1.9%)<br>7 (13.2%) | 2 (18.2%)<br>1 (9.1%)<br>0 (0%) | 0.483<br>0.316<br>0.339 |
| **RAP (mmHg)** | 8.9 ± 5.4 | 7.8 ± 5.8 | 0.574 |
| **mPAP (mmHg)** | 31.3 ± 10.8 | 24.7 ± 9.9 | 0.068 |
| **PAWP (mmHg)** | 18.6 ± 8.0 | 13.5 ± 6.7 | 0.041 |
| **HR (bpm)** | 73.7 ± 16.0 | 77.8 ± 14.9 | 0.426 |
| **Rhythm**<br>**Sinus Rhythm**<br>**Atrial Fibrillation/Flutter**<br>**Paced Rhythm**<br>**A-Paced**<br>**AV-Paced**<br>**V-Paced** | <br>30 (56.6%)<br>5 (9.4%)<br>18 (34.0%)<br>1 (1.9%)<br>6 (11.3%)<br>11 (20.8%) | <br>9 (81.8%)<br>1 (9.1%)<br>1 (9.1%)<br>0 (0%)<br>1 (9.1%)<br>0 (0%) | <br>0.178<br>1.000<br>0.151<br>1.000<br>1.000<br>0.187 |
| **SV (mL)** | 59.0 ± 17.2 | 91.4 ± 17.5 | < 0.001 |
| **CO (L/min)** | 4.1 ± 0.7 | 7.1 ± 0.8 | < 0.001 |
| **CI (L/min/m$^2$)** | 2.1 ± 0.4 | 3.2 ± 0.6 | < 0.001 |
