## Supplementary material for "Deep learning predicts cardiac output from seismocardiographic signals in heart failure": Central Illustration

**Central Illustration.** We describe a novel algorithm utilizing (1 and 2) publicly available wearable patch-derived electrocardiographic (ECG) and triaxial seismocardiographic (SCG) signals, combined with body mass index (BMI), in a (3) deep learning model to (4) predict cardiac output in heart failure.

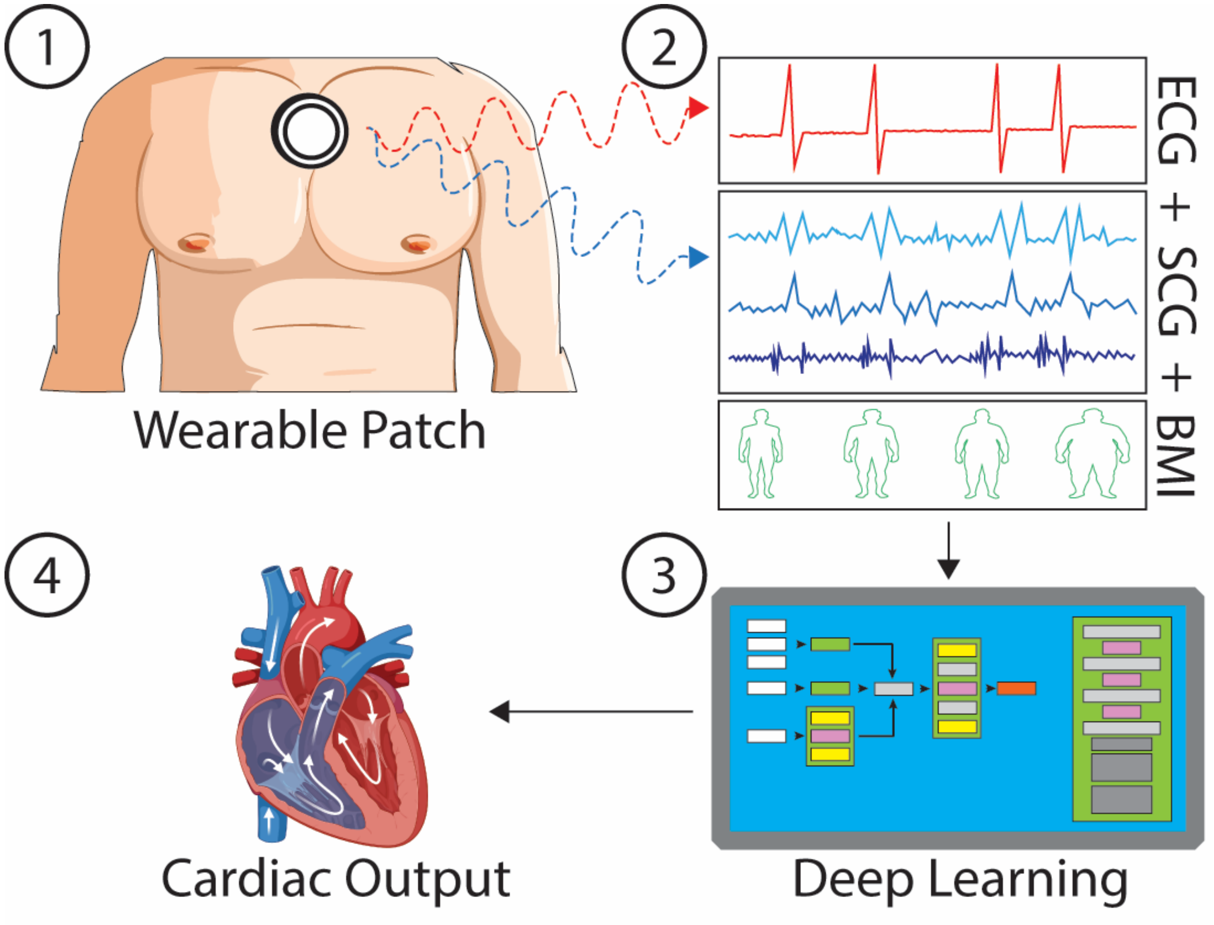
